# Early-life socioeconomic position and the accumulation of health-related deficits by midlife in the 1958 British birth cohort study

**DOI:** 10.1101/2020.09.14.20193961

**Authors:** Nina T Rogers, Joanna M Blodgett, Samuel D Searle, Rachel Cooper, Daniel H J Davis, Snehal M Pinto Pereira

**Author notes:** Corresponding author. UCL Division of Surgery & Interventional Science, University College London, London, UK.

## Abstract

Reducing population levels of frailty is an important goal and preventing its development in mid-adulthood could be pivotal. Childhood socioeconomic position (SEP) is associated with a myriad of adult health outcomes but evidence is limited on associations with frailty. Using 1958 British birth cohort data (N=8711), we aimed to: (i) establish the utility of measuring frailty in mid-life, by examining associations between a 34-item frailty index at 50y (FI_50y_) and mortality over an eight-year follow-up period and (ii) examine associations between early-life SEP and FI_50y_ and investigate whether these associations were explained by adult SEP. Hazard ratios (HRs) for mortality increased with increasing levels of frailty, e.g., HR_sex-adjusted_ was 4.07(95% CI:2.64,6.25) for highest vs. lowest fifth of FI_50y_. Lower early-life SEP was associated with higher FI_50y_: per unit decrease in early-life SEP (on a 4-point scale), FI_50y_ increased by 12.7%(10.85%,14.6%) in a model adjusted for early-life covariates. After additional adjustment for adult occupational class and education, the association attenuated to 5.71%(3.71%,7.70%). Findings suggest that early-life SEP is associated with frailty and that adult SEP only partially explains this association. Results highlight the importance of improving socioeconomic circumstances across the life course to reduce inequalities in frailty from mid-adulthood.

---

Frailty, a state of increased vulnerability as a consequence of age-related decline in physiological reserves[1], is associated with adverse health outcomes including falls, hospitalisations and premature mortality[1,2]. Therefore, frailty presents a global challenge because of population ageing[3,4]. Although prevalence of frailty increases with age, it is not limited to older ages[5]. Yet, most epidemiological studies assessing predictors of frailty have focused exclusively on adults aged 65 and over[6-8]. This omission is important because frailty reflects biological rather than chronological age[9] and is a dynamic process that may be reversible[10]. However, increasing age (from 65y onwards) is associated with a lower probability of improvement in frailty status[11]. Thus, there is emerging recognition that attention to frailty in mid-adulthood could be pivotal in terms of identifying, managing, and preventing severe frailty at older ages[12,13].

Reducing frailty at the population level is a desirable goal. To achieve this, a more precise understanding of predictors of frailty from mid-life onwards is key to delaying its onset. A life course approach to frailty has been discussed theoretically[14,15], and has the potential to identify when and how to intervene at different life-stages to maximize the chance of healthy population aging[14]. However, to date only few empirical life course studies have examined frailty. For example, a literature is emerging on links between early-life socioeconomic position (SEP) and frailty at older ages[8,16-20]. However, these studies have relied on relatively small sample sizes (N<1,100)[8,20], retrospective reporting of early-life SEP[16,17] and consideration of only a few other early-life covariates, such as birthweight, which have been shown to be associated with frailty[20]. Importantly, previous studies have examined mainly older adults and where younger adults have been considered[16,18-20], the age range has been broad, with little consideration for age-related differences in associations. While associations between frailty in adulthood and mortality are well established[1], evidence suggests that frailty levels may have increased in recent generations[21]. In addition, some[22] but not all[23] studies suggest that the strength of the frailty-mortality association may have weakened in more recent generations. Thus, there is utility in examining associations with mortality and predictive factors, such as early-life SEP, for frailty in a single-aged sample from mid-adulthood to help clarify when in the life course these associations emerge.

Despite the burgeoning literature linking early-life SEP to frailty, only a few studies [8,19,20] have examined whether this association is due to life course continuities in disadvantage. Limited evidence suggests that adult socioeconomic circumstances fully explain associations between early-life SEP and frailty at older ages[8,19,20]. However, no study has examined these chains of associations in mid-life or using a frailty index (FI). This index considers the accumulation of health-related deficits[24,25], and is a validated and commonly used approach to operationalising frailty. Compared with other frailty measures, the FI is more sensitive to small changes in health status[26] making it particularly suitable for examining frailty in mid-adulthood, a life-stage when health deficits are accumulating at a slower rate than at older ages [27].

We aim to address several outstanding research gaps regarding the utility of measuring frailty in mid-life and the links between early-life SEP and frailty. Specifically, using data from the 1958 British Birth Cohort, we derived a FI at 50y (referred to FI_50y_). To provide construct validity and establish the utility of measuring frailty in mid-life, we examined associations between FI_50y_ and mortality over an eight-year follow-up period. We then examined associations between early-life SEP and FI_50y_ and investigated whether these associations were explained by adult SEP.

## METHODS

The 1958 British Birth Cohort includes over 17,000 participants followed-up since birth during a single week in March 1958[28]. Ethical approval was given, including at 50y by the London multi-centre Research Ethics Committee and participants gave informed consent at various ages. Respondents in mid-adulthood are broadly representative of the surviving cohort[29]. At 50y, 9,789 individuals participated, of these 8,711 had a valid measure of FI_50y_ (see figure 1) and were included in the analysis. Compared to cohort members who took part at 50y, but had insufficient information to create a FI (N=1,078), participants included in this study had a more favorable SEP in early-life and in adulthood (Table SI).

**Figure 1:**
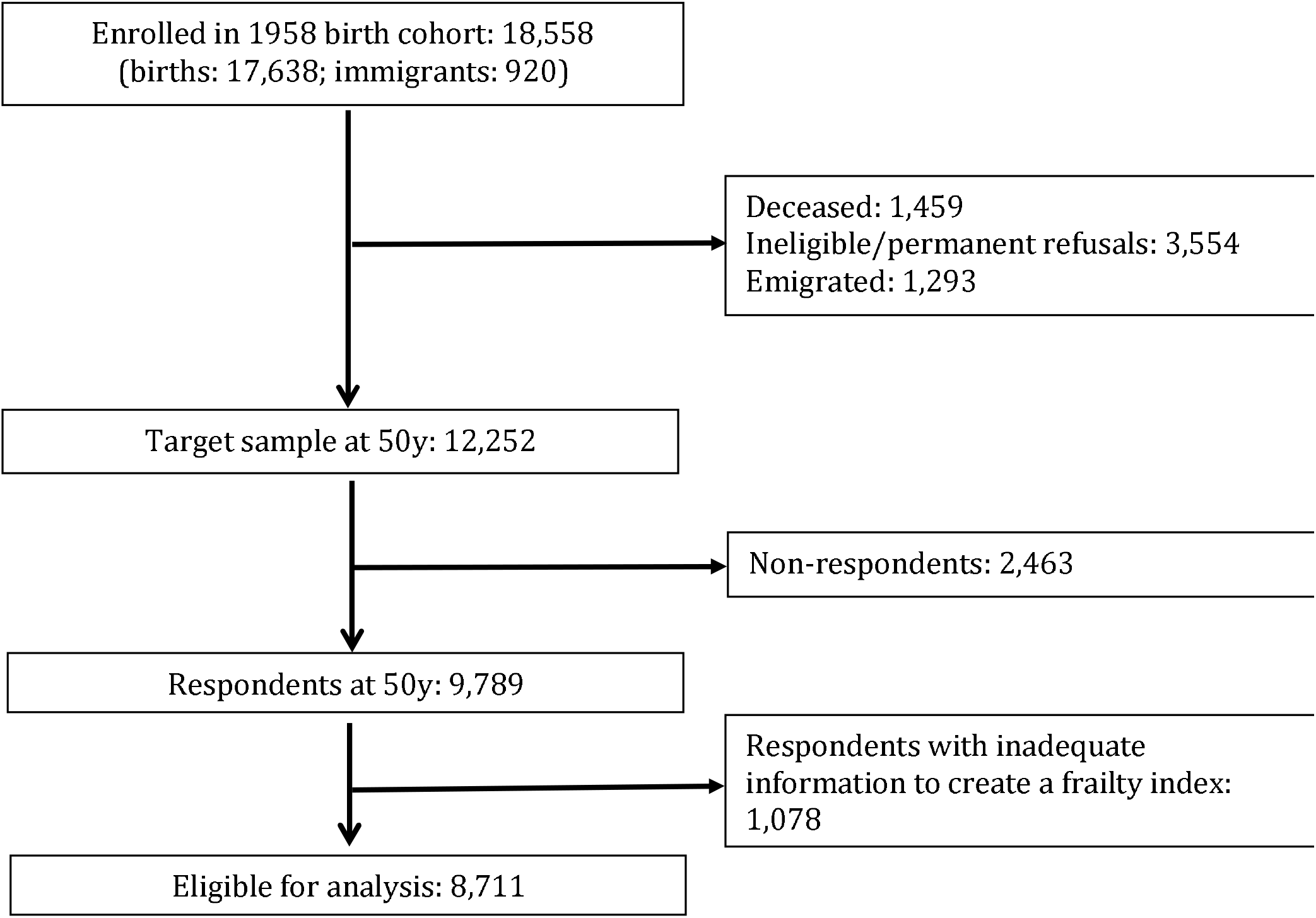
Flow diagram of participants from birth who were eligible to be included in analytical sample

*The frailty index:* was derived following standard guidelines [30]. Variables included in the index met the following criteria: a) a health-associated deficit with a prevalence that generally increases with age; b) not universal in the adult population by midlife (e.g. myopia is not included in the index but age-related sight changes (presbyopia) is included); and c) when taken together the included variables cover a range of physiological systems and processes. The FI_50y_ included 34 variables covering multiple physiological domains including chronic diseases, physical functioning and health, mental health, cognitive function, hearing and eyesight (see Table 1 for details). Most variables were dichotomised and given a score of 1 (deficit present) or 0 (deficit absent). Following published guidelines[30], individuals (N=8,711; 89.0%) were included provided they had information on at least 30 of the 34 deficits. For each included individual, FI_50y_ was generated by summing the total number of deficits reported and dividing this by the total number of deficits considered (i.e. number of considered deficits varied from 30 to 34), giving a continuous score between 0 and 1.

**Table 1:**
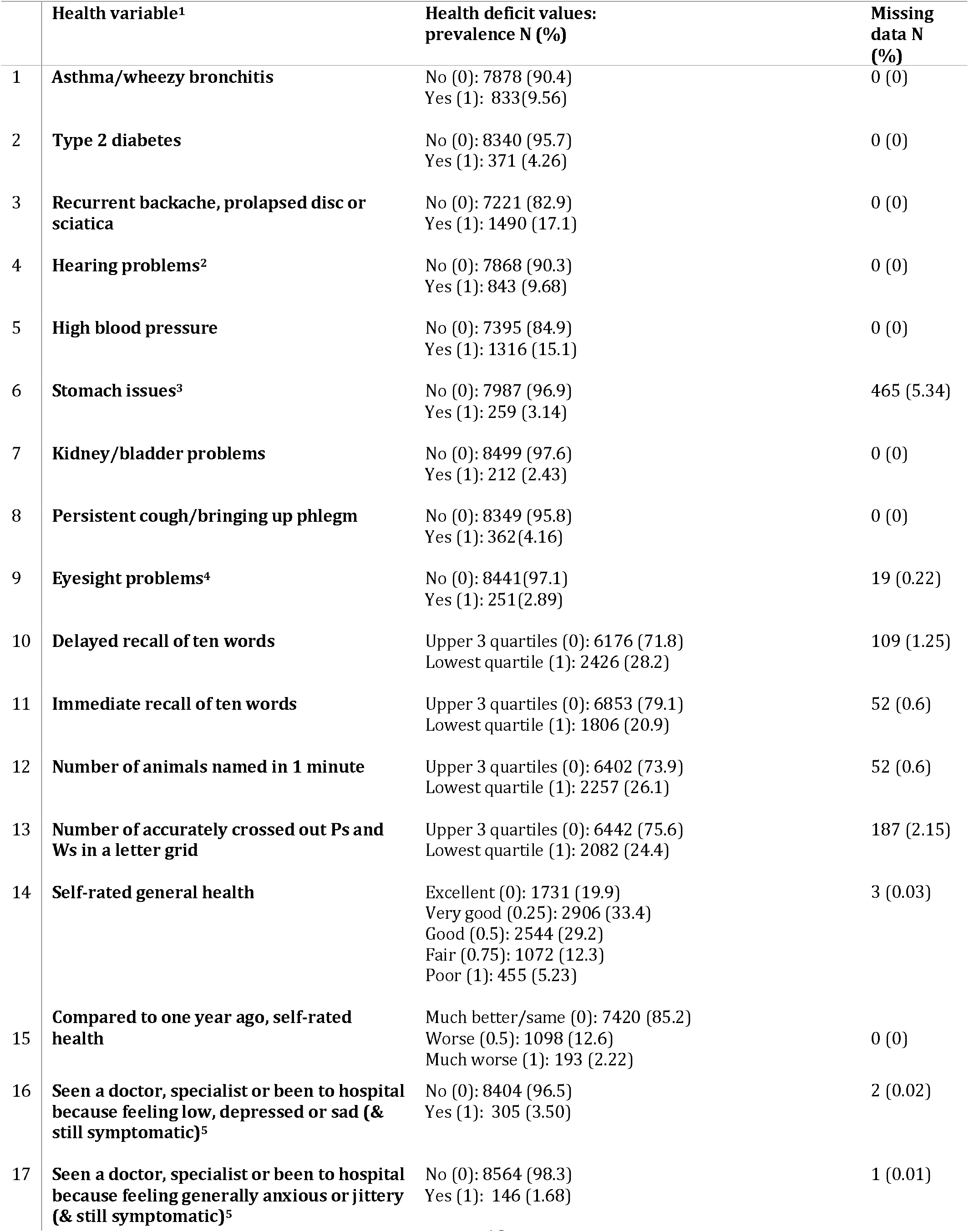
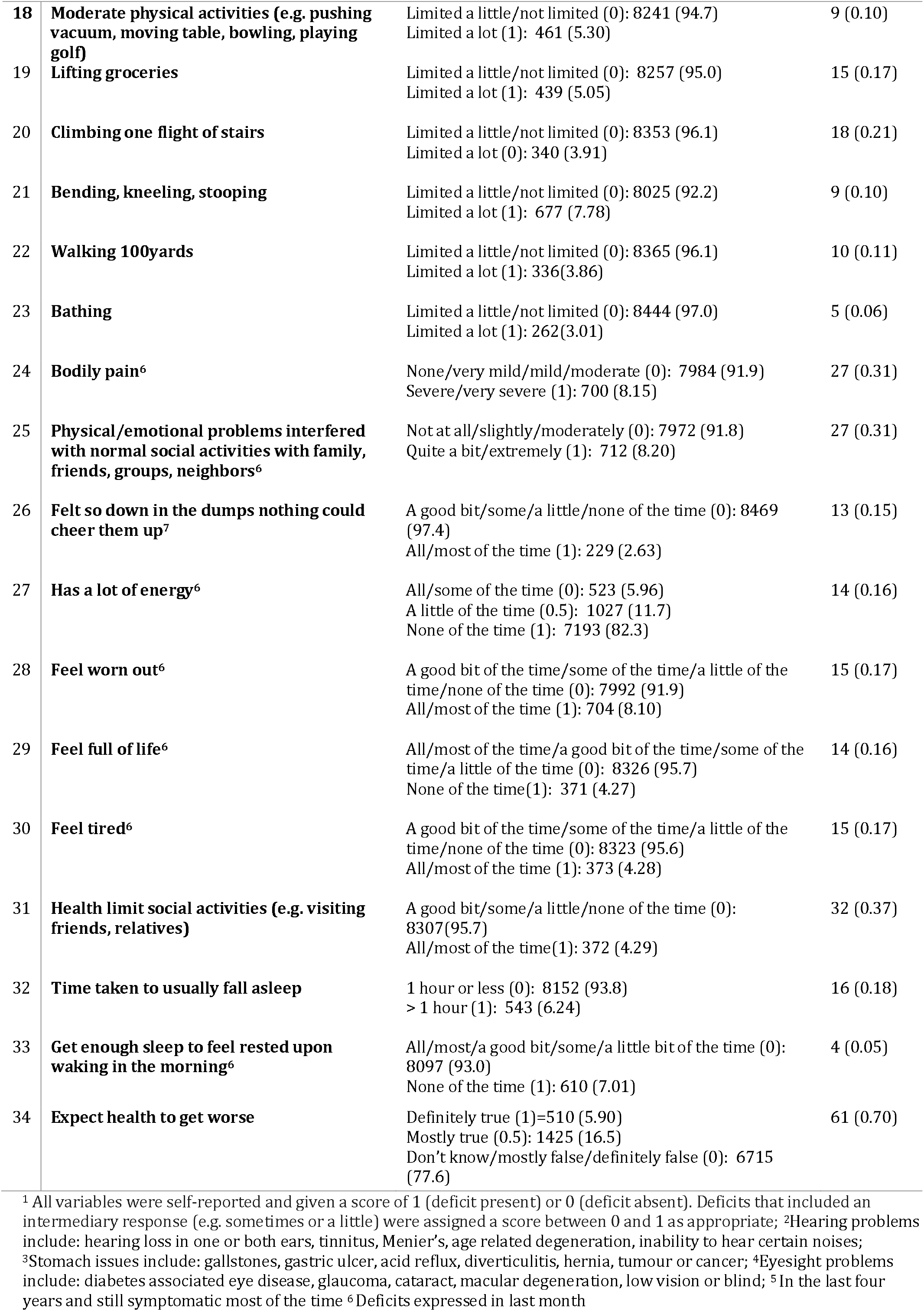
Health deficits used to construct the 34-item Frailty Index at 50y in the 1958 British birth cohort (N=8711)

|  | Health variable<sup>1</sup> | Health deficit values:<br>prevalence N (%) | Missing<br>data N<br>(%) |
| --- | --- | --- | --- |
| 1 | Asthma/wheezy bronchitis | No (0): 7878 (90.4)<br>Yes (1): 833(9.56) | 0 (0) |
| 2 | Type 2 diabetes | No (0): 8340 (95.7)<br>Yes (1): 371 (4.26) | 0 (0) |
| 3 | Recurrent backache, prolapsed disc or sciatica | No (0): 7221 (82.9)<br>Yes (1): 1490 (17.1) | 0 (0) |
| 4 | Hearing problems<sup>2</sup> | No (0): 7868 (90.3)<br>Yes (1): 843 (9.68) | 0 (0) |
| 5 | High blood pressure | No (0): 7395 (84.9)<br>Yes (1): 1316 (15.1) | 0 (0) |
| 6 | Stomach issues<sup>3</sup> | No (0): 7987 (96.9)<br>Yes (1): 259 (3.14) | 465 (5.34) |
| 7 | Kidney/bladder problems | No (0): 8499 (97.6)<br>Yes (1): 212 (2.43) | 0 (0) |
| 8 | Persistent cough/bringing up phlegm | No (0): 8349 (95.8)<br>Yes (1): 362(4.16) | 0 (0) |
| 9 | Eyesight problems<sup>4</sup> | No (0): 8441(97.1)<br>Yes (1): 251(2.89) | 19 (0.22) |
| 10 | Delayed recall of ten words | Upper 3 quartiles (0): 6176 (71.8)<br>Lowest quartile (1): 2426 (28.2) | 109 (1.25) |
| 11 | Immediate recall of ten words | Upper 3 quartiles (0): 6853 (79.1)<br>Lowest quartile (1): 1806 (20.9) | 52 (0.6) |
| 12 | Number of animals named in 1 minute | Upper 3 quartiles (0): 6402 (73.9)<br>Lowest quartile (1): 2257 (26.1) | 52 (0.6) |
| 13 | Number of accurately crossed out Ps and Ws in a letter grid | Upper 3 quartiles (0): 6442 (75.6)<br>Lowest quartile (1): 2082 (24.4) | 187 (2.15) |
| 14 | Self-rated general health | Excellent (0): 1731 (19.9)<br>Very good (0.25): 2906 (33.4)<br>Good (0.5): 2544 (29.2)<br>Fair (0.75): 1072 (12.3)<br>Poor (1): 455 (5.23) | 3 (0.03) |
| 15 | Compared to one year ago, self-rated health | Much better/same (0): 7420 (85.2)<br>Worse (0.5): 1098 (12.6)<br>Much worse (1): 193 (2.22) | 0 (0) |
| 16 | Seen a doctor, specialist or been to hospital because feeling low, depressed or sad (& still symptomatic)<sup>5</sup> | No (0): 8404 (96.5)<br>Yes (1): 305 (3.50) | 2 (0.02) |
| 17 | Seen a doctor, specialist or been to hospital because feeling generally anxious or jittery (& still symptomatic)<sup>5</sup> | No (0): 8564 (98.3)<br>Yes (1): 146 (1.68) | 1 (0.01) |
| 18 | **Moderate physical activities (e.g. pushing vacuum, moving table, bowling, playing golf)** | Limited a little/not limited (0): 8241 (94.7)<br>Limited a lot (1): 461 (5.30) | 9 (0.10) |
| 19 | **Lifting groceries** | Limited a little/not limited (0): 8257 (95.0)<br>Limited a lot (1): 439 (5.05) | 15 (0.17) |
| 20 | **Climbing one flight of stairs** | Limited a little/not limited (0): 8353 (96.1)<br>Limited a lot (0): 340 (3.91) | 18 (0.21) |
| 21 | **Bending, kneeling, stooping** | Limited a little/not limited (0): 8025 (92.2)<br>Limited a lot (1): 677 (7.78) | 9 (0.10) |
| 22 | **Walking 100yards** | Limited a little/not limited (0): 8365 (96.1)<br>Limited a lot (1): 336(3.86) | 10 (0.11) |
| 23 | **Bathing** | Limited a little/not limited (0): 8444 (97.0)<br>Limited a lot (1): 262(3.01) | 5 (0.06) |
| 24 | **Bodily pain<sup>6</sup>** | None/very mild/mild/moderate (0): 7984 (91.9)<br>Severe/very severe (1): 700 (8.15) | 27 (0.31) |
| 25 | **Physical/emotional problems interfered with normal social activities with family, friends, groups, neighbors<sup>6</sup>** | Not at all/slightly/moderately (0): 7972 (91.8)<br>Quite a bit/extremely (1): 712 (8.20) | 27 (0.31) |
| 26 | **Felt so down in the dumps nothing could cheer them up<sup>7</sup>** | A good bit/some/a little/none of the time (0): 8469 (97.4)<br>All/most of the time (1): 229 (2.63) | 13 (0.15) |
| 27 | **Has a lot of energy<sup>6</sup>** | All/some of the time (0): 523 (5.96)<br>A little of the time (0.5): 1027 (11.7)<br>None of the time (1): 7193 (82.3) | 14 (0.16) |
| 28 | **Feel worn out<sup>6</sup>** | A good bit of the time/some of the time/a little of the time/none of the time (0): 7992 (91.9)<br>All/most of the time (1): 704 (8.10) | 15 (0.17) |
| 29 | **Feel full of life<sup>6</sup>** | All/most of the time/a good bit of the time/some of the time/a little of the time (0): 8326 (95.7)<br>None of the time(1): 371 (4.27) | 14 (0.16) |
| 30 | **Feel tired<sup>6</sup>** | A good bit of the time/some of the time/a little of the time/none of the time (0): 8323 (95.6)<br>All/most of the time (1): 373 (4.28) | 15 (0.17) |
| 31 | **Health limit social activities (e.g. visiting friends, relatives)** | A good bit/some/a little/none of the time (0): 8307(95.7)<br>All/most of the time(1): 372 (4.29) | 32 (0.37) |
| 32 | **Time taken to usually fall asleep** | 1 hour or less (0): 8152 (93.8)<br>> 1 hour (1): 543 (6.24) | 16 (0.18) |
| 33 | **Get enough sleep to feel rested upon waking in the morning<sup>6</sup>** | All/most/a good bit/some/a little bit of the time (0): 8097 (93.0)<br>None of the time (1): 610 (7.01) | 4 (0.05) |
| 34 | **Expect health to get worse** | Definitely true (1)=510 (5.90)<br>Mostly true (0.5): 1425 (16.5)<br>Don't know/mostly false/definitely false (0): 6715 (77.6) | 61 (0.70) |

<sup>1</sup> All variables were self-reported and given a score of 1 (deficit present) or 0 (deficit absent). Deficits that included an intermediary response (e.g. sometimes or a little) were assigned a score between 0 and 1 as appropriate; <sup>2</sup>Hearing problems include: hearing loss in one or both ears, tinnitus, Menier's, age related degeneration, inability to hear certain noises; <sup>3</sup>Stomach issues include: gallstones, gastric ulcer, acid reflux, diverticulitis, hernia, tumour or cancer; <sup>4</sup>Eyesight problems include: diabetes associated eye disease, glaucoma, cataract, macular degeneration, low vision or blind; <sup>5</sup> In the last four years and still symptomatic most of the time <sup>6</sup> Deficits expressed in last month

*Mortality:* Information on deaths from 2008 (when cohort members were 50y) to the end of 2016 (when cohort members were 58y) was ascertained from a variety of sources, the majority (94.7%; n=198) through linkage to death certificates from the National Health Service Central Register[31]. Information from relatives or close friends during survey activities/cohort maintenance allowed identification of 11 further deaths (details in Table 3 footnotes).

*Early-life Socioeconomic Position:* was identified from prospectively recorded information on father’s occupation at birth in 1958 or if missing at 7yin 1965 (n=631 (7.24%)). Using the Registrar General’s Social Classification groupings, four categories were identified: professional/managerial (classes I and II), skilled non-manual (class III non-manual), skilled manual (class III manual) and partly skilled/unskilled manual (classes IV and V and cases in which there was no male head of household).

### Covariates and potential intermediaries

Covariates were selected a-priori following review of the literature [8,32] and included sex, maternal smoking during pregnancy, maternal age at birth, birthweight (adjusted for gestational age), breastfeeding status (<1 month; ≥1 month) and birth order. All factors were reported by parents, except birthweight, which was ascertained from clinical records. Adult SEP was considered a potential intermediary based on established associations with both early-life SEP[33,34] and frailty[21,35]. It was represented here by occupational class at42y (or if missing at 33y (n=829(9.52%)), grouped into four categories from professional/managerial (classes I and II) to partly skilled/unskilled manual (classes IV and V) and educational attainment by 33y, grouped into four categories from <0-levels to degree or higher (see Table 2 footnotes for details).

#### Statistical analysis

Proportional hazards for mortality were visually assessed using Kaplan-Meier plots. Cox proportional hazard models (sex-adjusted) estimated hazard ratios and 95% confidence intervals (HR(95%CI)) of associations between the FI_50y_and all-cause mortality between ages 50y-58y. Associations between FI and mortality are commonly examined using a continuous measure [36] or FI categories derived from specific cut-points (e.g. 0-0.1, 0.1-0.2, etc) [2,37,38]. However, in mid-life, the FI is highly skewed (e.g. ~60% of the sample have a FI<0.1). Thus these categorizations are not appropriate. Therefore, similar to other studies examining associations between the FI and mortality, we divided FI_50y_ into fifths [39]. Survival time included time from completion of the 50y survey to date of death, censoring (last date of contact) or end of the study period (December 2016), whichever came first Schoenfeld residuals were checked to test the assumption of proportional hazards for FI_50y_ and sex; neither violated the assumption. We examined associations between early-life SEP and FI_50y_ using linear regression models. For ease of interpretation, FI_50y_ was log-transformed and multiplied by 100, whereby the regression coefficients can be interpreted as the symmetric percentage difference in means[40]. Before log-transforming FI_50y_, we added 0.01 to the index, as in previous work[41], to circumvent logarithm values of zero. In these models, we first adjusted for early-life covariates listed above and then to assess the role of adult SEP as a potential intermediary, we further adjusted for adult occupational class and education. To determine whether early-life and adult SEP acted synergistically, we examined interactions between early-life and adult occupational class (dichotomized into non-manual and manual categories); there was no evidence of interaction (*P*=0.74). We examined whether associations for early-life SEP varied by sex. There was no evidence of effect modification (*P*=0.35), hence sex-adjusted analyses are presented. In supplementary analyses, we explored the influence of adult occupational class and educational attainment separately. Missing data ranged from 5.0% (for maternal age at birth) to 13.9% (gestational age). To minimize data loss, missing information for covariates, early-life and adult SEP were imputed using multiple imputation-chained equations. Following published recommendations[42], imputation models included all substantive variables and main predictors of missingness (age 7-year internalizing and externalizing behaviours and cognitive ability) [29]. Analyses were run across 20 imputed datasets and overall estimates were obtained. Imputed results were similar to those obtained using observed values; the former are presented.

## RESULTS

Social mobility between early-life and adulthood was substantial: while 20% of the cohort had fathers in professional/managerial occupations when they were born, 42% of the cohort were themselves in professional/managerial occupations at 42y (Table 2). As expected, the FI_50y_ was right skewed, with a median of 0.07 for both males and females, corresponding to an expression of approximately 2 (34*0.07) health-related deficits.

**Table 2:** Characteristics of the 1958 British birth cohort (N=8711)^a^

|  | Total population | Females | Males |
| --- | --- | --- | --- |
| Early-life SEP<sup>b</sup> |  |  |  |
| Father's occupational class |  |  |  |
| I/II | 1671 (19.7) | 856 (19.5) | 815 (20.0) |
| III non-manual | 877 (10.36) | 439 (9.98) | 438 (10.8) |
| III manual | 4054 (47.9) | 2110(48.0) | 1944 (47.8) |
| IV/V | 1865 (22.0) | 994(22.6) | 871 (21.4) |
| Adult occupational class<sup>b</sup> |  |  |  |
| I/II | 3368 (42.1) | 1530 (36.9) | 1838 (47.8) |
| III non-manual | 1836 (23.0) | 1449(35.0) | 387 (10.1) |
| III manual | 1475 (18.5) | 288 (6.95) | 1187 (30.8) |
| IV/V | 1313 (16.4) | 876 (21.1) | 437 (11.4) |
| Adult education<sup>c</sup> |  |  |  |
| <O-levels | 1596 (21.4) | 944 (23.8) | 652 (18.6) |
| O-levels | 2548 (34.1) | 1503 (37.9) | 1045 (29.8) |
| A-levels | 2245 (30.1) | 1013 (25.6) | 1232 (35.1) |
| Degree or higher | 1083 (14.5) | 505 (12.7) | 578 (16.5) |
| Frailty Index: Median (Q25, Q75) | 0.07 (0.04,0.13) | 0.07 (0.03,0.13) | 0.07 (0.04,0.13) |

<sup>a</sup> Based on observed (i.e. unimputed) data
<sup>b</sup> Early-life SEP based on father's occupation at birth (or if missing at 7y); adult occupational class at 42y (or if missing at 33y). Both classified using Register General's classification of occupation and grouped into: professional/managerial (classes I and II), skilled non-manual (class III non-manual), skilled manual (class III manual) and partly/unskilled manual (classes IV and V; in early-life also included cases where there was no male head of household).
<sup>c</sup> Educational attainment based on highest educational qualification by 33y. O-levels: high school qualifications typically ascertained at age 16y. A-levels: National qualifications typically ascertained at age 18y

Sex-adjusted Kaplan-Meier curves indicated that mortality generally increased progressively with increasing levels of frailty (figure 2). For example,compared to the least frail adults (i.e. those in the lowest frailty fifth), the sex-adjusted HR was 1.66(1.01,2.74) for adults in the fourth highest frailty fifth and 4.07(2.64,6.25) for the most frail adults (i.e. those in the highest frailty fifth, Table S2).

**Figure 2:**
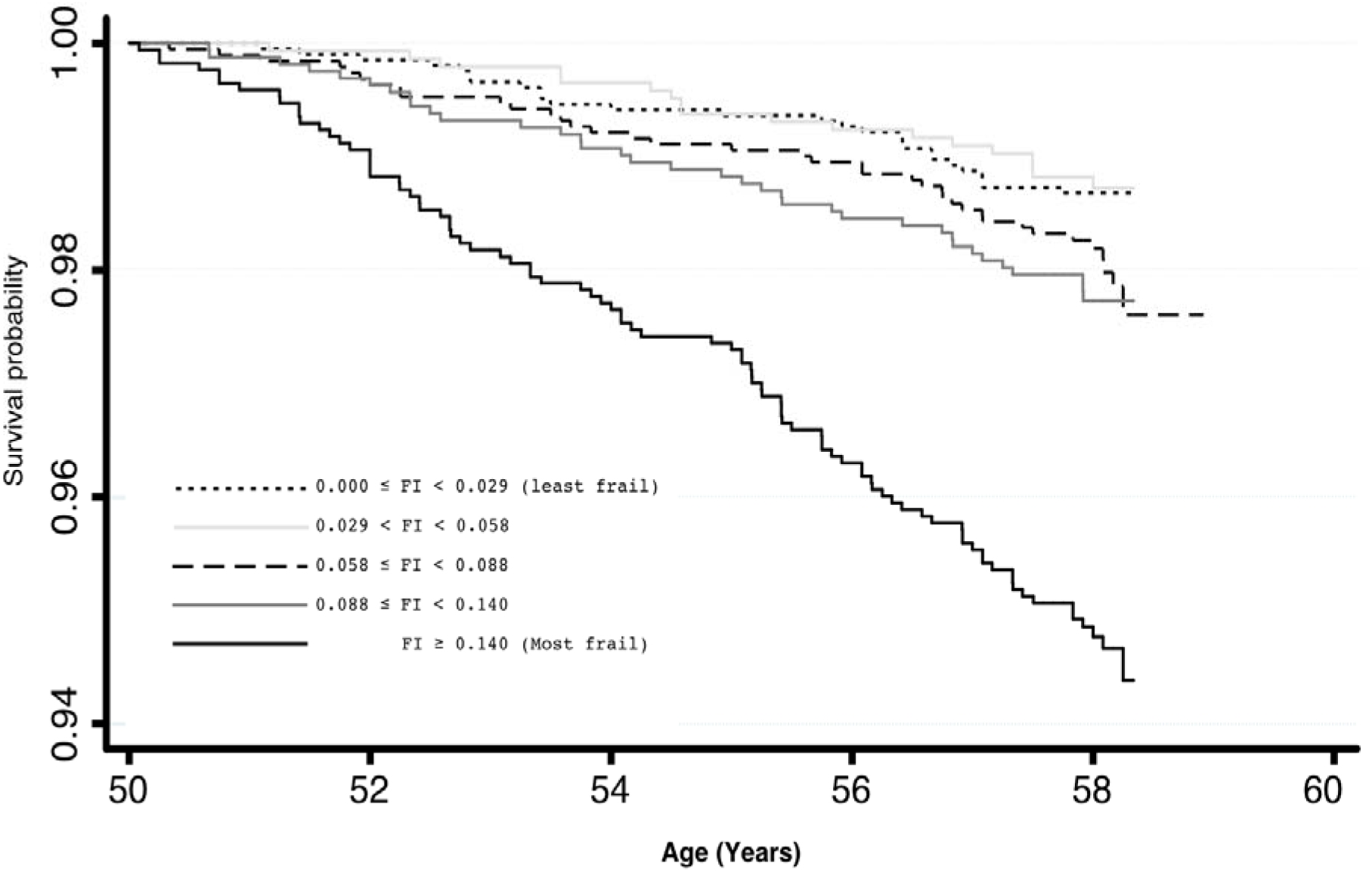
Sex-adjusted Kaplan Meier survival probabilities stratified by frailty fifths in the 1958 birth cohort (n=8711, 209 deaths)

Lower early-life SEP was associated with greater percentage differences in FI_50y_. For example, after adjusting for covariates, FI_50y_ was higher by 8.54% (1.16%, 15.9%) for participants with fathers in III non-manual, 25.7% (20.5%, 30.8%) for III manual and 36.8% (30.7%, 43.0%) for IV/V when compared to participants with fathers in class I/II (Table 3, model 2). Associations attenuated but remained after adjustment for adult occupational class and education. For example, associations were reduced to 10.3% (5.15%, 15.5%) for participants with fathers in III manual and 16.1% (9.90%,22.4%) for IV/V (Table 3, model 3). Separate adjustment for adult SEP and education in turn, showed that both had attenuating effects (Table S3).

**Table 3:**
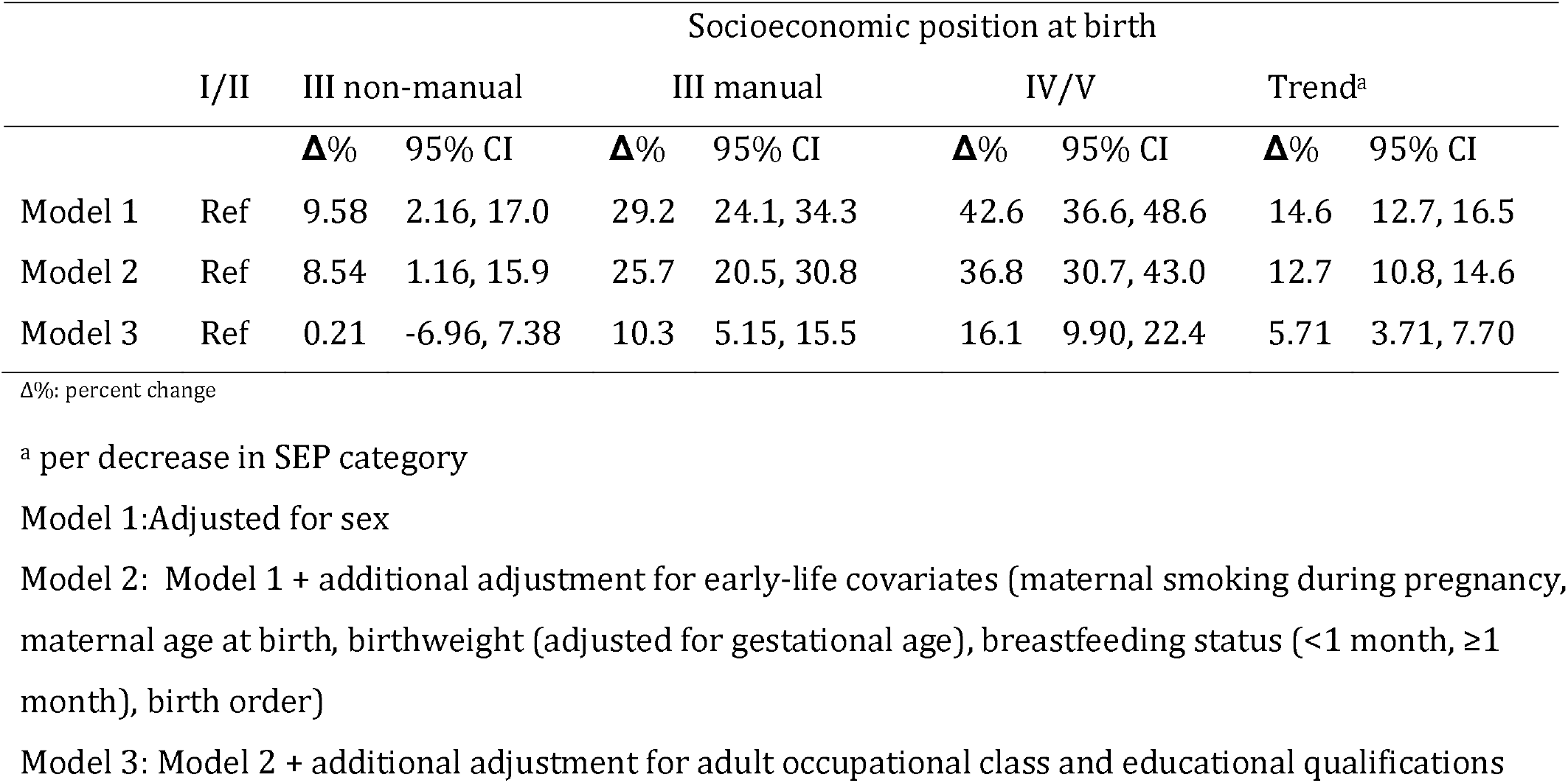
Mean percentage difference (95% confidence interval) in frailty index at 50y by socioeconomic position at birth

## DISCUSSION

Our study examining early-life SEP in relation to the accumulation of health-related deficits by mid-adulthood, in a general population sample, is important for several reasons. First, we show that by mid-life, health deficits have begun to accumulate. For example, a median FI of 0.07 indicates that half the population at 50y had at least two of the considered health deficits and similarly, a quarter of the population had at least four deficits. Second, it was noteworthy that this accumulation of deficits at a relatively young age was strongly associated with mortality up to eight years later. For example, hazards of mortality were over four times higher, comparing adults with the most (≥5) to those with the least (0-1) number of deficits. Third, lower SEP in early-life was associated with higher levels of frailty by mid-adulthood, such that the FI for those born in the lowest SEP category was over 36% greater at 50y compared to those born in the highest SEP category. Finally, the link between early-life SEP and the accumulation of health deficits by mid-life was partly explained by continuities in disadvantage into adulthood.

A major study strengths is the examination of an age-homogenous sample, which has been lacking in other studies. This is advantageous since the influence of age, which is associated strongly with frailty, can be eliminated when examining associations between early-life SEP and frailty in this study. Further strengths include examination of a large general population sample with prospective data from birth to adulthood, a validated measure of frailty capturing multiple physiological domains and the consideration of several important early-life covariates, such as maternal smoking during pregnancy, maternal age at birth, birthweight, breastfeeding status and birth order. Examining socioeconomic circumstances at just one life-stage is likely to be inadequate to fully elucidate the contribution of SEP at different life-stages for subsequent health risks[43]. Thus, examining socioeconomic circumstances at two distinct phases across the life course is another study strength. We acknowledge that there is no single best indicator of SEP[44]. We used father’s occupation at birth to represent early-life SEP because it is a commonly used measure, reflecting a wide range of early-life social and economic indicators including the household’s educational attainment, income levels and social standing. In addition, rather than using a single measure of SEP in adulthood we used two well-established indicators (educational attainment and occupational class). Health deficits accumulate at a slower rate in mid-life than at older ages [27] and frailty measured in younger populations might be clinically and biologically different from that measured in older populations[1]. Nonetheless, our measure of frailty is particularly suited to mid-life because it has demonstrated good construct validity at this life stage[45] and it provides a continuous score of fitness to frail [24] allowing detection of small differences in health compared to other measures[26]. Finally, as with all long-term studies, attrition occurred over time. Although participants in this study had more favourable early-life and adult SEP compared to those not included at 50y, in general the sample remains broadly representative of the original cohort[29]. Further sample reductions due to missing data were prevented using multiple imputation following published guidelines[42].

Our findings that health deficits have already begun to accumulate by 50y and predict subsequent mortality agrees with the literature on the accumulation of health deficits in mid-life. For example, our measure of frailty at 50y is broadly in agreement with the few other studies examining the FI ata similar life-stage[37,46]. Although the implications of frailty in clinical practice may vary by age, we and others[13,46] demonstrate the utility of measuring frailty earlier in the life course. Thus, our findings emphasise that measuring frailty at a particular age is meaningful in identifying individuals at risk of adverse health outcomes and, because frailty is progressive, beginning with a preclinical stage, there are opportunities for early prevention[1].

Our results are consistent with previous studies showing that lower early-life SEP is associated with greater risk of frailty in adulthood[8,19,20]. We found that associations were maintained after controlling for a broad range of prospectively measured early-life covariates such that each reduction in SEP category in early-life (on a four-point scale) was associated with approximately a 13% lower FI at 50y. Associations attenuated, but remained, after adjustment for adult SEP. This is noteworthy because, as argued elsewhere [47], associations for adult SEP might partly be due to health-related social mobility, whereas those for early-life SEP cannot be. Therefore our findings suggest that while influences on mid-life frailty are found in both childhood and adulthood, life-time SEP appears to not act synergistically in relation to mid-life frailty, instead a cumulative effects life course model is more likely[48]. Since adult SEP did not fully explain early-life SEP associations in this cohort, other explanatory pathways may also be involved. Evidence from the literature suggests that early-life socioeconomic disadvantage may lead to poor adult health via biological embedding[49]. For example, abnormal biological changes have been observed in adults who experienced early-life socioeconomic disadvantage in this cohort[47] and elsewhere[50] which have been proposed to lead to accelerated ageing[51]. In contrast to our findings, in previous studies, adult SEP fully explained early-life SEP—frailty associations. Discrepancies may be due to the single-aged sample examined here compared with the broad age ranges previously examined[19,20] or the younger age of adults in this study compared to others[8]. Notwithstanding this difference, our findings are in agreement with others, that adult SEP is an important intermediary through which early-life SEP is associated with midlife frailty. Growing up in socioeconomically disadvantaged circumstances is predictive of poor socioeconomic outcomes (e.g. low educational achievement) in adulthood[52] which in turn, is linked to frailty[53]. Therefore, our findings, together with other evidence, suggests that interventions to improve adult socioeconomic circumstances of those from disadvantaged backgrounds may reduce the burden of frailty in mid-life and beyond.

In conclusion, our findings have several practical and policy relevant implications. They emphasize the value of using previously collected health data to identify those who may be vulnerable to accelerated ageing earlier in the life course. Derivations of the FI are already widely used in clinical and primary care settings in England[24,54] to systematically identify the extent of frailty in adults aged 65 and over. Our findings suggest that similar assessments could be valuable in mid-adulthood and suggests that in a primary care setting, in addition to considering single health deficits in mid-life, the accumulation of deficits is also important Identifying adults in mid-life who could benefit from early interventions might reduce the burden of frailty at older ages, improving quality of life and reducing costs of care[13,46]. We highlight the importance of improving socioeconomic conditions over the whole life course in order to reduce health inequalities. Thus, a potential intervention focus could be on improving socioeconomic opportunities available in adulthood for those disadvantaged in childhood. Moreover, relative child poverty is projected to rise from 29.7% to 36.6% in the UK between 2018 to 2022 [5 5], thus our findings underscore the importance of much needed policies to redress socioeconomic inequalities in childhood because they have the potential to improve health in mid-adult life and beyond.

### Contributors

NTR and SMPP conceived the study and drafted the paper. JB and SS advised on construction of the frailty index at 50y. NTR carried out the analysis. All authors contributed to the interpretation of data, revision of the manuscript, and approved its final version.

## Data Availability

All data are available on registration at the UK Data Service. The authors are grateful to the Centre for Longitudinal Studies (CLS), UCL Institute of Education, for the use of the 1958 cohort data and to the UK Data Service for making them available. However, neither CLS nor the UK Data Service bear any responsibility for the analysis or interpretation of these data.

https://discover.ukdataservice.ac.uk/series/?sn=2000032

## Acknowledgments

Application for all-cause mortality data was made through the UK Data service. The authors are grateful to the Centre for Longitudinal Studies (CLS), UCL Institute of Education, for the use of the 1958 cohort data and to the UK Data Service for making them available. However, neither CLS nor the UK Data Service bear any responsibility for the analysis or interpretation of these data.

## Funding

This work was supported by a UK Medical Research Council Career Development Award (ref: MR/P020372/1) to SMPP. The views expressed in the publication are those of the authors and not necessarily those of the funders. The funders had no input into study design; data collection, analysis, and interpretation; in the writing of the report; and in the decision to submit the article for publication. Researchers were independent of influence from study funders.

**Table S1:** Prevalencea of early-life and adult socioeconomic position in the included and not included cohort at 50y

|  | Included (n=8711) | Not included (n=1078) |
| --- | --- | --- |
| Early-life SEP<sup>b</sup> |  |  |
| Father's occupational class |  |  |
| I/II | 1671 (19.7) | 128 (12.5) |
| III non-manual | 877 (10.36) | 82 (7.99) |
| III manual | 4054 (47.9) | 549 (53.5) |
| IV/V | 1865 (22.0) | 267 (26.0) |
| Adult occupational class<sup>b</sup> |  |  |
| I/II | 3368 (42.1) | 326 (36.3) |
| III non-manual | 1836 (23.0) | 179 (19.9) |
| III manual | 1475 (18.5) | 222 (24.7) |
| IV/V | 1313 (16.4) | 172 (19.1) |
| Adult education<sup>c</sup> |  |  |
| <O-levels | 1596 (21.4) | 229 (28.0) |
| O-levels | 2548 (34.1) | 282 (34.5) |
| A-levels | 2245 (30.1) | 224 (27.4) |
| Degree or higher | 1083 (14.5) | 83 (10.2) |

<sup>a</sup> Table based on observed (i.e. unimputed) data
<sup>b</sup> Early-life SEP based on father's occupation at birth (or if missing at 7y); adult occupational class at 42y (or if missing at 33y). Both classified using Register General's classification of occupation and grouped into: professional/managerial (classes I and II), skilled non-manual (class III non-manual), skilled manual (class III manual) and partly/unskilled manual (classes IV and V; in early-life also included cases where there was no male head of household).
<sup>c</sup> Educational attainment based on highest educational qualification by 33y. O-levels: high school qualifications typically ascertained at age 16y. A-levels: National qualifications typically ascertained at age 18y.

**Table S2:**
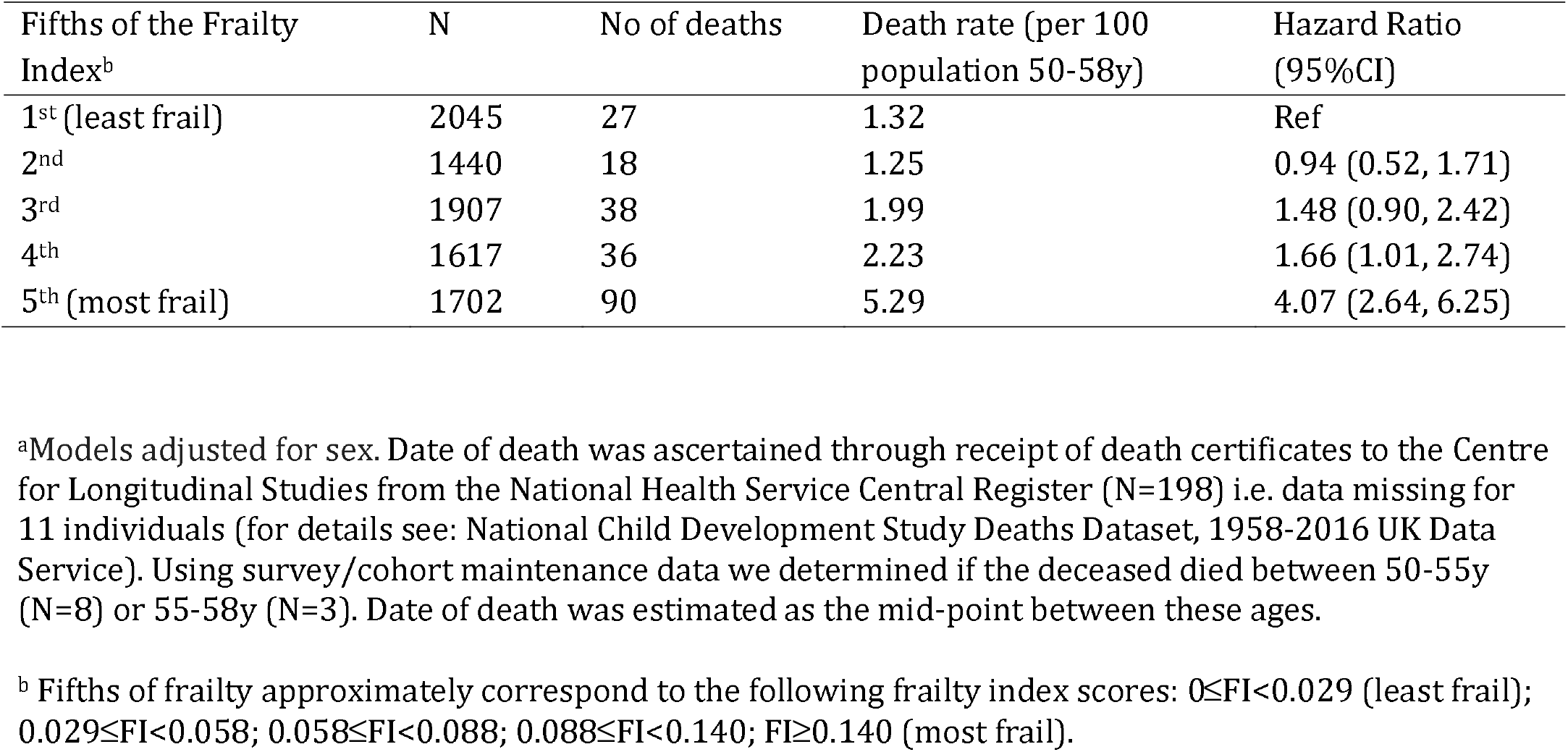
Hazard ratios (95% confidence intervals) for fifths of the frailty index at 50y in relation to all-cause mortality in 1958 birth cohort participants aged 50y to 58y^a^ (N=8711)

| Fifths of the Frailty Index<sup>b</sup> | N | No of deaths | Death rate (per 100 population 50-58y) | Hazard Ratio (95%CI) |
| --- | --- | --- | --- | --- |
| 1<sup>st</sup> (least frail) | 2045 | 27 | 1.32 | Ref |
| 2<sup>nd</sup> | 1440 | 18 | 1.25 | 0.94 (0.52, 1.71) |
| 3<sup>rd</sup> | 1907 | 38 | 1.99 | 1.48 (0.90, 2.42) |
| 4<sup>th</sup> | 1617 | 36 | 2.23 | 1.66 (1.01, 2.74) |
| 5<sup>th</sup> (most frail) | 1702 | 90 | 5.29 | 4.07 (2.64, 6.25) |

<sup>a</sup>Models adjusted for sex. Date of death was ascertained through receipt of death certificates to the Centre for Longitudinal Studies from the National Health Service Central Register (N=198) i.e. data missing for 11 individuals (for details see: National Child Development Study Deaths Dataset, 1958-2016 UK Data Service). Using survey/cohort maintenance data we determined if the deceased died between 50-55y (N=8) or 55-58y (N=3). Date of death was estimated as the mid-point between these ages.
<sup>b</sup> Fifths of frailty approximately correspond to the following frailty index scores: $0 \leq FI < 0.029$ (least frail); $0.029 \leq FI < 0.058$ ; $0.058 \leq FI < 0.088$ ; $0.088 \leq FI < 0.140$ ; $FI \geq 0.140$ (most frail).

**Table S3:**
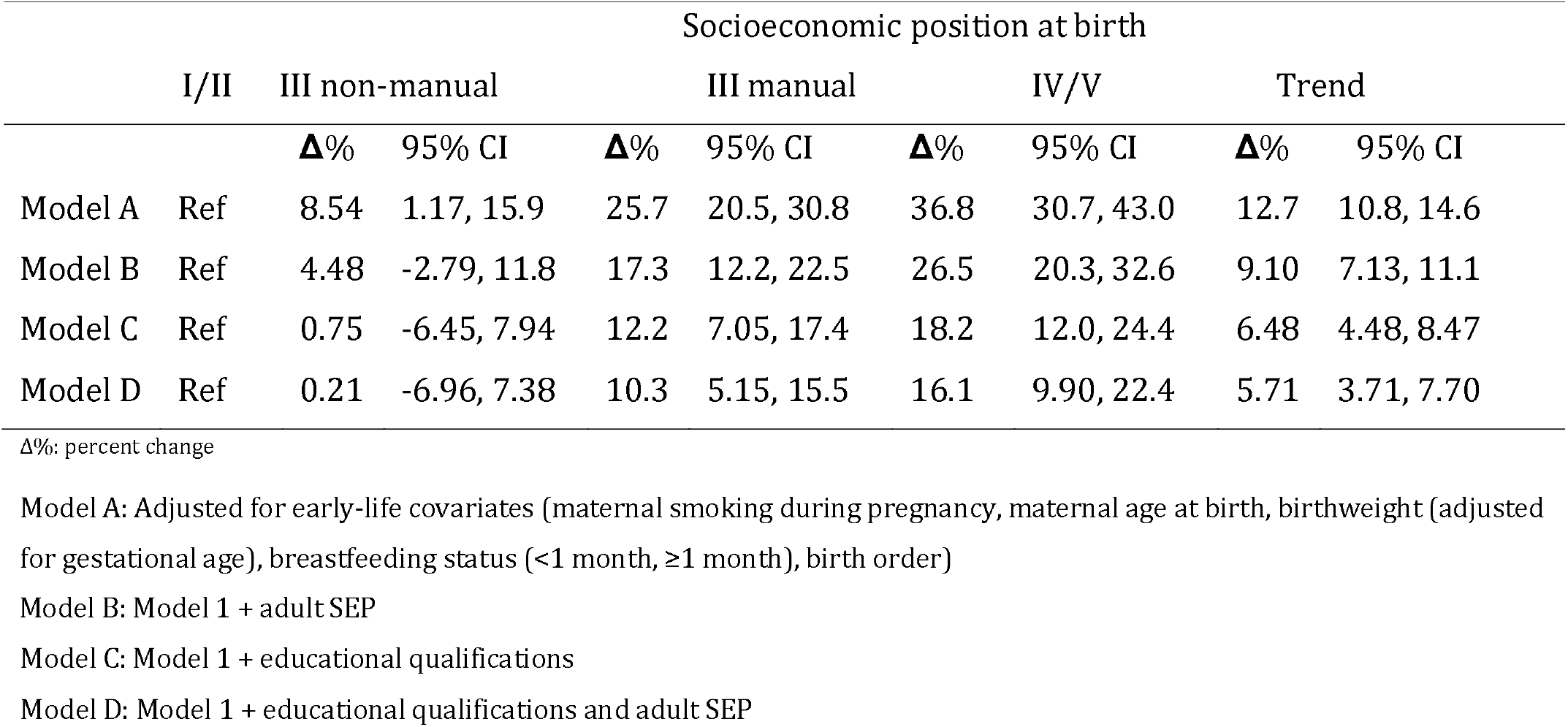
Mean percentage difference (95% confidence interval) in FI50y by socioeconomic position at birth considering adult occupational class (Model B) and educational attainment (Model C) separately

## Abbreviations

FI (Frailty Index); FI_50y_ (Frailty Index at 50 years); SEP (socioeconomic position)

